# Vaccines alone are no silver bullets: a modeling study on the impact of efficient contact tracing on COVID-19 infection and transmission

**DOI:** 10.1101/2021.08.29.21262789

**Authors:** Dhesi Baha Raja, Nur Asheila Abdul Taib, Alvin Kuo Jing Teo, Vivek Jason Jayaraj, Choo-Yee Ting

**Affiliations:** Ainqa Health, Malaysia; Saw Swee Hock School of Public Health, National University of Singapore, National University Health System, Singapore; Department of Social and Preventive Medicine, Faculty of Medicine, Universiti Malaya, Malaysia; Faculty of Computing and Informatics, Multimedia University, Malaysia

**Author notes:** Corresponding author: Choo-Yee Ting, Address: Faculty of Computing and Informatics, Multimedia University, Persiaran Multimedia, 63100 Cyberjaya, Selangor, Malaysia.

## Abstract

**Background:** The computer simulation presented in this study aimed to investigate the effect of contact tracing on COVID-19 transmission and infection in the context of rising vaccination rates.

**Methods:** This study proposed a deterministic SEIRV model with contact tracing and vaccination components. We initialized some parameters using the Malaysian COVID-19 data to inform the model. We defined contact tracing effectiveness as the proportion of contacts of a positive case that was successfully traced and vaccination rate as the proportion of daily doses administered per population in Malaysia. Sensitivity analyses on the untraced and infectious populations were conducted. The study presented in silico findings on multiple scenarios by varying the contact tracing effectiveness and daily vaccination rates.

**Results:** At a vaccination rate of 1.4%, a contact tracing with the effectiveness of 70% could delay the peak of untraced asymptomatic cases by 17 days and reduce the highest number of daily cases by 70% compared with a 30% contact tracing effectiveness. A similar trend was observed for symptomatic cases when a similar experiment setting was used. We also performed sensitivity analyses by using different combinations of contact tracing effectiveness and vaccination rates. In all scenarios, the effect of contact tracing on COVID-19 incidence persisted for both asymptomatic and symptomatic cases.

**Conclusion:** Despite testing only on two public health and social measures (PHSMs), we observed the scenario with low contact tracing and increasing vaccination rates successfully mimicked the current transmission trend in Malaysia. Hence, while vaccines are progressively rolled out, efficient contact tracing must be rapidly implemented concurrently to reach, find, test, isolate, and support the affected populations to bring the pandemic under control.

## Introduction

The pandemic caused by the virus SARS-CoV-2 has infected more than 200 million people and led to 4.5 million deaths worldwide as of 26 August 2021.^1^ Since January 2021, approximately 2 billion people are fully vaccinated against COVID-19, with several countries, mostly upper-middle-income economies, having reached 70% full vaccination to-date.^1,2^ Despite this, the pandemic shows no sign of abatement, with the spread of the *Delta* variant triggering new outbreaks in many countries globally.^3^

Governments of various countries have started to invest in digital contract tracing since 2020. Digital contact tracing is an approach to disrupt the chains of disease transmission by identifying and isolating those in close contact with an infected individual.^4^ It leverages proximity and geospatial technologies to provide a comprehensive approach to collect spatio-temporal data.^5,6^ The data can be used to study the movement and interaction of humans across time, which can then be utilized further for investigating disease transmissions such as COVID-19, tuberculosis, and other communicable diseases. A successful digital contact tracing could keep the reproduction number R-naught under control, while failing to have an efficient digital contract tracing would cause the daily cases to increase exponentially. A study by Mizumoto and colleagues has shown that for COVID-19, 70% of transmissions occur before someone is symptomatic,^7^ indicating the importance of having a speedy and accurate contract tracing mechanism. Another study by Abueg and colleagues has also reported that if a digital contact tracing is used by 75% of the population, the number of infections can be reduced by 73–79%.^8^

Among those fully vaccinated, reports of breakthrough infections, severe illness, and deaths have since been reported in countries like Iran and Indonesia.^9,10^ The evidence on the protective effect of several COVID-19 vaccines in the World Health Organization Emergency Use Listing against the Delta variant has gradually emerged, with their effectiveness reported being lower than the protection conferred against the Alpha variant.^11–13^ Nevertheless, the evidence thus far indicates that vaccines are effective against symptomatic and severe COVID-19,^12,14,15^ and vaccines uptake and administration should be ramped up globally. From a public health perspective, it is vital to reduce the transmission and incidence of infection to protect pockets of populations who could not be vaccinated and allow economies to open in a safe and calibrated manner.

As of 26 August 2021, the cumulative number of COVID-19 cases in Malaysia has exceeded 1.5 million, and the daily new confirmed cases per 100,000 population remain one of the highest in the world.^1^ While Malaysia has consistently rolled out between 400,000 and 500,000 doses of vaccines per day since July 2021 and implemented multiple iterations of movement control order with different measures since March 2020, the pandemic continues to rage with a record number of cases and deaths daily. Contact tracing efforts are also severely hampered due to the strain to the public health system, causing missed contacts who might have been infected but did not know their risk to delay testing and further transmit the virus.^16,17^

It is increasingly evident that a single intervention, be it vaccine, public health, or social measures, is insufficient to control the pandemic. Hence, in this paper, we aimed to investigate the importance of implementing an effective contact tracing on COVID-19 transmission and infection in the context of rising vaccination rates using a deterministic, compartmental modeling approach as an experimental basis for our discussion.

## Methods

### Design

First, we proposed a novel transmission model which factored in contact tracing effectiveness and vaccination. Next, we determined the parameter using estimations based on Malaysia COVID-19 data and information from published literature. Finally, we conducted sensitivity analyses of the parameters on the number of untraced, infectious individuals.

### Epidemic model

We developed a deterministic, compartmental SEIRV model to study the transmission dynamics of COVID-19 when contact tracing and vaccination were incorporated. The human population was subdivided into ten classes according to their disease status, namely the susceptible (*S*), exposed (*E*), traced exposed (*T*), quarantined symptomatic infected (*Q*^*sym*^), quarantined asymptomatic infected (*Q*^*asym*^), symptomatic infected (*I*^*sym*^), asymptomatic infected (*I*^*asym*^), recovered (*R*), death (*D*) and vaccinated (*V*). The susceptible compartment (*S*) was composed of all healthy individuals who could get infected with SARS-CoV-2. Individuals in the exposed compartment (*E*) were those who have gotten infected with the virus but remained in their latent period and untraced. The traced exposed (*T*) compartment referred to infectives in their latent period who were successfully traced and isolated. Quarantined symptomatic infected (*Q*^*sym*^) compartment comprised infected individuals who were infectious with symptoms and in quarantine either at home or in hospital, whereas the quarantined asymptomatic infected (*Q*^*asym*^) compartment was composed of those infectious individuals without symptoms and in quarantine. Individuals in the symptomatic infected (*I*^*sym*^) compartment referred to those untraced infectious individuals who have developed the symptoms, while those untraced infectious individuals without symptoms belonged in the asymptomatic infected (*I*^*asym*^) compartment. Those who recovered with COVID-19 immunity made up the recovered (*R*) compartment, and those who received COVID-19 immunity through vaccination formed the vaccinated (*V*) compartment. Lastly, victims who died from COVID-19 were represented by the compartment *D*. Individuals could transition from one compartment over time but were only allowed to be in one compartment at a time. These compartments were summarized in Table 1.

**Table 1:**
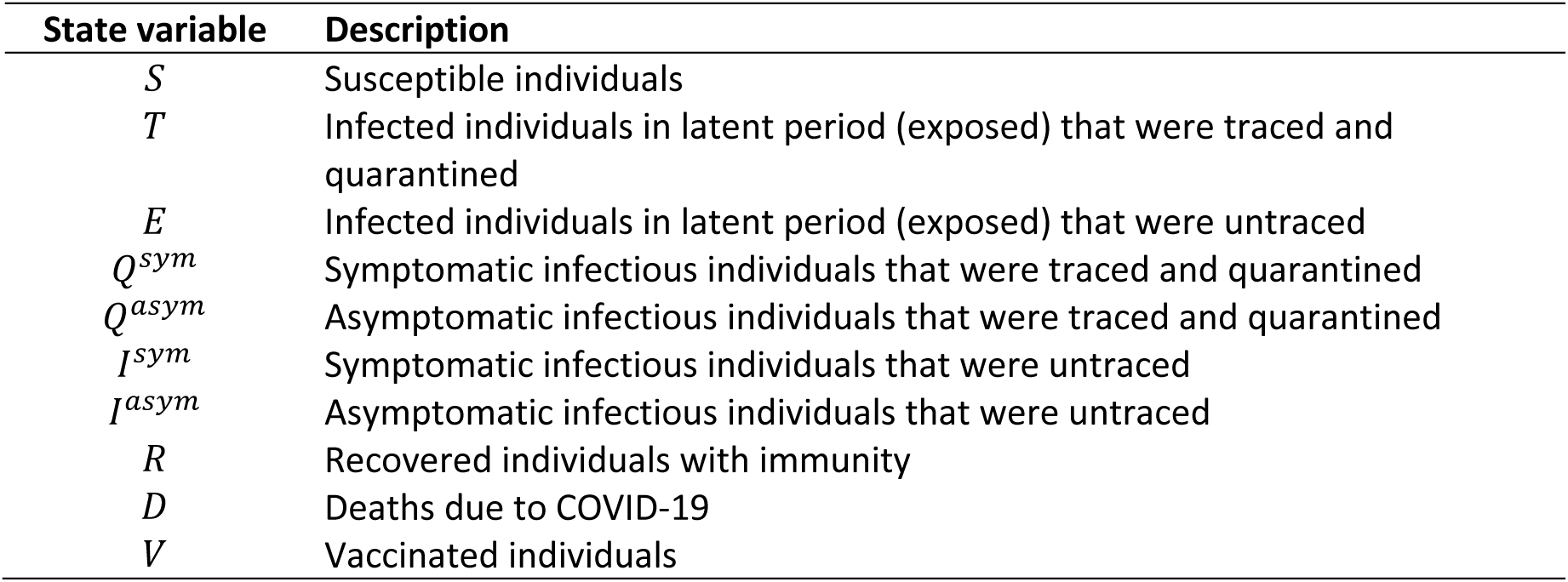
Description of compartments in our SEIRV model

### Model assumptions

The susceptibles (*S*) could become infectives when they met with either symptomatic (*I*^*sym*^) or asymptomatic (*I*^*asym*^) infectious individuals at different transmission rates of *β*_*sym*_ or *β*_*asym*_, respectively. This transmission rate was the product of contact rate and the probability of transmission given contact. In this paper, we assumed that these public health and social measures (PHSM) did not vary across time and that the population was not partitioned according to age or comorbidity. Also, natural births and deaths were not considered. Our focus was on analyzing the “trace and isolate” policy, whereby tracing could be done manually or through an automated process using tracing apps. Hence, we subdivided the exposed compartment (infectives in latent period) further into traced exposed (*T*) and untraced exposed (*E*) compartments. The implementation of the “trace and isolate” approach could reduce the transmission of COVID-19 by forcing the traced exposed individuals into quarantine (*T*) through self-isolation. Therefore, we assumed that all the traced individuals in quarantine (*T*) will have full compliance.

As they were unaware of their disease status, asymptomatic infectious individuals (*I*^*asym*^) would continue to contribute to the transmission of the virus when they met another susceptible at a rate of *β*_*asym*_, leading to untraced infectives in the *E* compartment. Following the work of Grimm and colleagues,^18^ the parameter *ω* denoted the proportion of the symptomatic infectious population (*I*^*sym*^) detected by the health authorities, while the parameter *τ* was the contact tracing effectiveness which described the fraction of the contacts traced either manually or via digital tools such as tracing apps. Hence, the product *ωτ* gave the total proportion of infectives in their latent period who were successfully traced and transitioned into the (*T*) compartment at a rate of *β*_*sym*_, whereas (1 - *ωτ*) referred to tracing failures and thus, (1 - *ωτ*) proportion of infectives would enter the untraced exposed (*E*) compartment at a rate of *β*_*sym*_.

Furthermore, infectives in their latent period (*E, T*) would become infectious at a rate of *σ*, which denoted the reciprocal of the incubation period. With *ε* as the fraction of exposed persons (*E, T*) who were asymptomatic, then *εσE* would be the number of untraced exposed (*E*) entering the asymptomatic infectious compartment (*I*^*asym*^), while *εσT* referred to the number of traced exposed (*T*) who would move into the asymptomatic infectious quarantine (*Q*^*asym*^) compartment. This gives (1 - *ε*) as the fraction of the exposed population (*E, T*) who were symptomatic, which leads to (1 - *ε*)*σE*, the number of untraced exposed (*E*) moving into the symptomatic infectious compartment (*I*^*sym*^), whereas (1 - *ε*)*σT* as the number of traced exposed (*T*) entering the asymptomatic infectious quarantine (*Q*^*sym*^) compartment. We assumed that only symptomatic infected individuals could die from COVID-19 at a rate of *μ* thus entering the *D* compartment. We also assumed that the disease-induced death rate (*μ*) was constant and unaffected by disease severity and hospital capacity. Symptomatic infected individuals (*I*^*sym*^, *Q*^*sym*^) would recover at a rate of *γ*_*sym*_ which was the reciprocal of the duration of infectiousness of symptomatic patients. On the other hand, asymptomatic infected individuals (*I*^*asym*^, *Q*^*asym*^) would recover at a rate of *γ*_*asym*_ which was the reciprocal of the duration of infectiousness of asymptomatic patients. Finally, we incorporated vaccination into the model. In this work, the vaccine functioned by reducing the number of susceptibles, thus preventing further infections at a rate of *vp*, which referred to the product of vaccination coverage and effectiveness. The parameters (1 - *p*) and *α* denoted the probability of vaccination failure in preventing transmissions and the reciprocal duration for the waning of vaccine immunity, respectively. Hence, (1 - *p*)*αV* gave the number of vaccinated individuals who would move back to being susceptible (*S*). The dynamics of transmission were visualized in Figure 1 with the description of parameters listed in Table 2:

**Figure 1:**
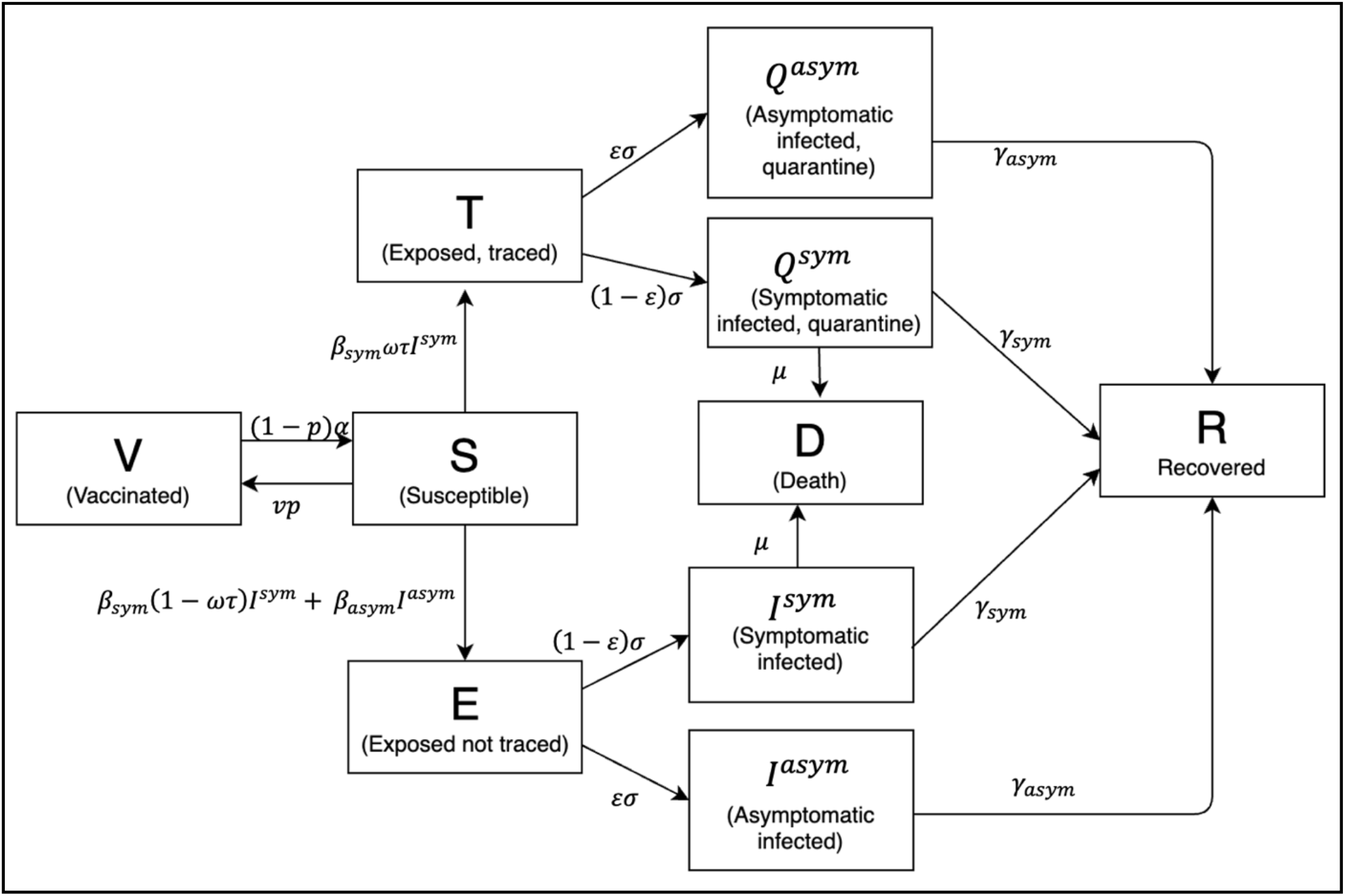
Transmission diagram of the SEIRV model.

**Table 2:**
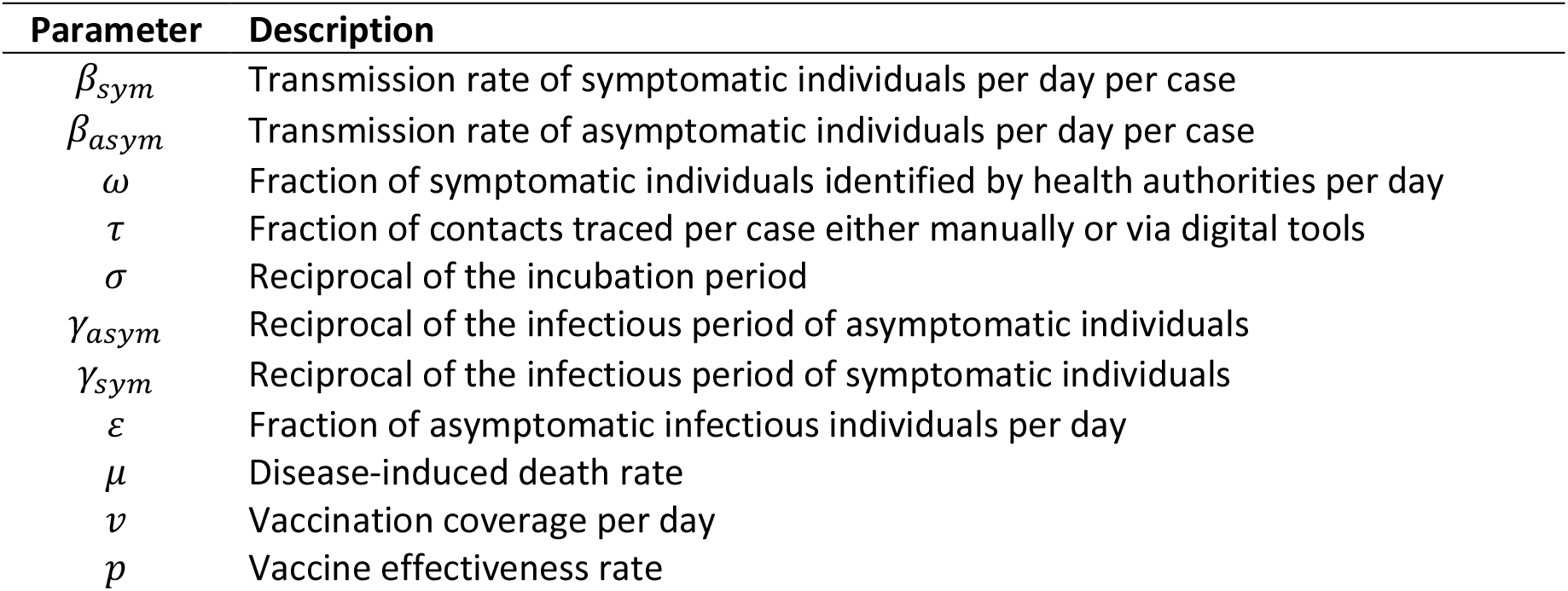
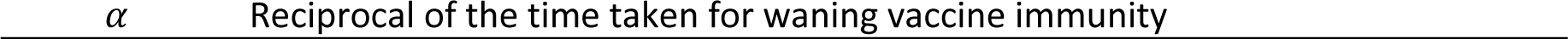
Parameter description

| Parameter | Description |
| --- | --- |
| $\beta_{sym}$ | Transmission rate of symptomatic individuals per day per case |
| $\beta_{asym}$ | Transmission rate of asymptomatic individuals per day per case |
| $\omega$ | Fraction of symptomatic individuals identified by health authorities per day |
| $\tau$ | Fraction of contacts traced per case either manually or via digital tools |
| $\sigma$ | Reciprocal of the incubation period |
| $\gamma_{asym}$ | Reciprocal of the infectious period of asymptomatic individuals |
| $\gamma_{sym}$ | Reciprocal of the infectious period of symptomatic individuals |
| $\varepsilon$ | Fraction of asymptomatic infectious individuals per day |
| $\mu$ | Disease-induced death rate |
| $v$ | Vaccination coverage per day |
| $p$ | Vaccine effectiveness rate |

### Model equations

Following the previous assumptions, our susceptible-exposed-infected-vaccinated (SEIRV) model was described by a nonlinear system of ten ordinary differential equations with 12 parameters:

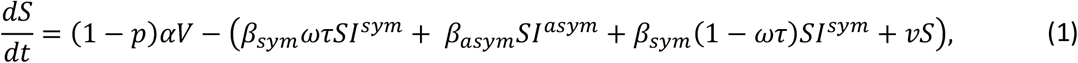

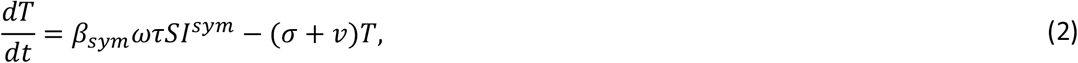

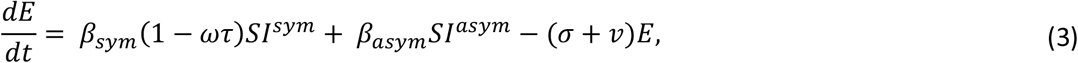

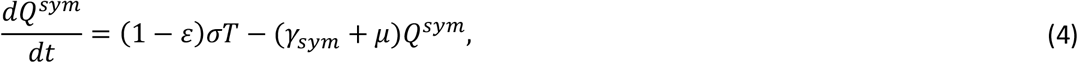

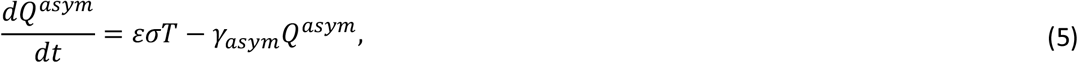

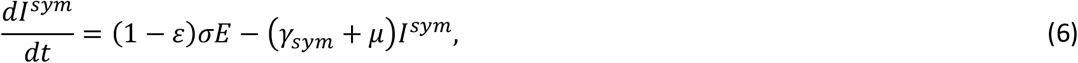

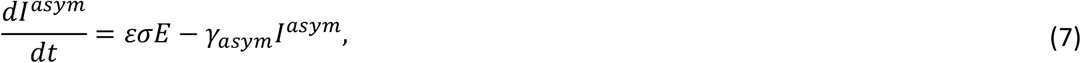

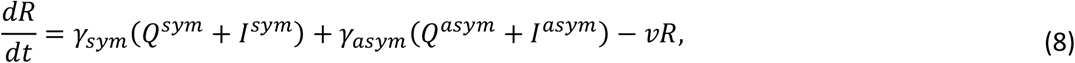

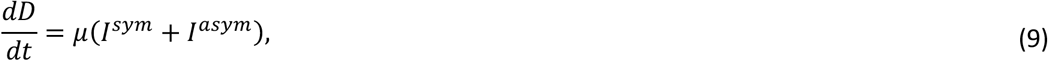

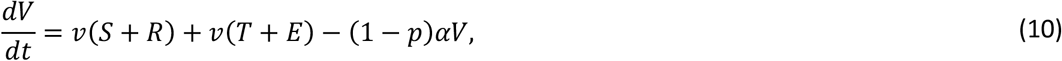

with initial conditions *S*(0) = *S*_0_ > 0, *T* > 0, *E*(0) = *E*_0_ > 0, *Q*^*sym*^ (0) = *Q*^*sym*^_0_ > 0, *Q*^*asym*^ (0) = *Q*^*asym*^_0_ > 0, *I*^*sym*^ (0) = *I*^*sym*^_0_ > 0, *I*^*asym*^ (0) = *I*^*asym*^_0_ > 0, *R*(0) = *R*_0_ > 0, *D*(0) = *D*_0_ > 0, and *V*(0) = *V*_0_ > 0.

### Parameter estimation

The values of parameters in this paper were either estimated using Malaysian COVID-19 data or adapted from literature. From a COVID-19 modeling study in Malaysia by Gill and colleagues,^19^ we took the average contacts per day per case (n=25) and the probability of transmission given contact by symptomatic persons = 0.05. We calculated the transmission rate *β* as the product of the two which gave us 25 × 0.05 = 1.25. However, since we assumed that the population was closed with constant size N = 32600000 (the total Malaysian population), we divided the transmission rate *β* by N, to get our final *β*_*sym*_ as 3.8 × 10^−8^. To calculate *β*_*asym*_, we followed the same steps but replaced the probability of transmission given contact by asymptomatic persons = 0.02 from Churches and colleagues,^20^ to get the *β*_*asym*_ as 1.5 × 10^−8^.

We defined the effectiveness of contact tracing as the proportion of contacts of a positive case that was successfully traced. In Malaysia, we estimated that the proportion of contacts of a COVID-19 case traced per case varied between 30% to 40%.^21,22^ The vaccination coverage per day (*ν*) was estimated from the Malaysian vaccination statistics, where around 1.0%–1.5% of the total population was vaccinated daily. Also, we included the values for the vaccination coverage, *ν* = 0.8% and the contact tracing effectiveness, *τ* = 50%, 70%, 90% as well for our simulation exercise. Other parameters obtained from the literature review were summarized in Table 3.

**Table 3:** Parameter values

| Parameter | Value | Source |
| --- | --- | --- |
| $\beta_{sym}$ | $3.8 \times 10^{-8}$ | 19 |
| $\beta_{asym}$ | $1.5 \times 10^{-8}$ | 20 |
| $\omega$ | 0.9 | 19 |
| $\tau$ | 0.9,0.7,0.5,0.4,0.3 | estimated, dynamic |
| $\sigma$ | 1/5 | 18 |
| $\gamma_{asym}$ | 1/10 | 18 |
| $\gamma_{sym}$ | 1/12.5 | 18 |
| $\varepsilon$ | 0.25 | 18 |
| $\mu$ | 0.02 | 20 |
| $v$ | 0.014,0.012,0.01,0.008 | estimated, dynamic |
| $p$ | 0.9 | 23 |
| $\alpha$ | 1/30 | 23 |

### Sensitivity analysis

In order to solve our SEIRV model, we implemented a numerical integration method Runge-Kutta of order 5 by using *solve_ivp* function from the *scipy*.*integrate* module in Python along with parameter values in Table 3. Next, to study the dynamics of transmission of COVID-19 concerning contact tracing and vaccination, we conducted sensitivity analyses of the parameters on the untraced, infectious individuals. We were interested in observing the simulation on *I*^*asym*^ and *I*^*sym*^ because these populations would contribute to forward transmission since they were not traced and isolated. Hence, an uncontrolled number of *I*^*asym*^ and *I*^*sym*^ would in turn lead to a potential surge in future total COVID-19 cases. We varied the values of vaccination coverage *ν* and contact tracing effectiveness *τ* and investigated the effects of the changes on *I*^*asym*^ and *I*^*sym*^. We prepared four scenarios as summarized in Table 4:

**Table 4:** Scenarios for sensitivity analysis

| Scenario | Fixed parameter | Value (/day) | Varying parameter | Value (/day) |
| --- | --- | --- | --- | --- |
| 1 | Vaccination coverage | 1.4% | Contact tracing effectiveness | 30%, 40%, 50%, 70%, 90% |
| 2 |  |  | (Vaccination coverage, Contact tracing effectiveness) | (30%, 1.5%), (40%, 1.4%), (50%, 1.3%), (70%, 1.2%), (90%, 1%) |
| 3 | Contact tracing effectiveness | 90% | Vaccination coverage | 0.8%, 1%, 1.3%, 1.4% |
| 4 | Contact tracing effectiveness | 35% | Vaccination coverage | 0.8%, 1%, 1.3%, 1.4% |

## Results

Scenario 1 simulated five different contact tracing effectiveness ranged between 30% and 90% against the backdrop of a fixed vaccination rate of 1.4% per day (**Figure 2a-2b**). This scenario assumed that the vaccine was administered at a rate of approximately 450,000 doses per day. We found that when contact tracing effectiveness was at 30%, the number of untraced, asymptomatic cases would peak at day-42 with approximately 1.52 million cases before gradually tapering down—the peak decreases as the contact tracing effectiveness increases (**Figure 2a**). When contact tracing effectiveness was increased to 70%, the peak was delayed by about 17 days, with the highest number of daily cases at 459,000, which was about a 70% reduction from those estimated when contact tracing effectiveness was at 30%. It can be observed that a contact tracing effectiveness of 90% would almost flatten the curve. Similar behavior could be observed for the untraced, symptomatic cases in **Figure 2b**, in which a combination of high vaccination rate (1.4%) and low contact tracing effectiveness (30%) would cause a peak of the cases to be about 3.5 million at day-40. When we increased the contact tracing effectiveness to 70%, the peak was delayed by about 15 days, with approximately one million cases which were about 29% of those estimated when contact tracing was at 30% effectiveness. When contact tracing effectiveness increased, the peaks for both cases were delayed and lowered.

**Figure 2a:**
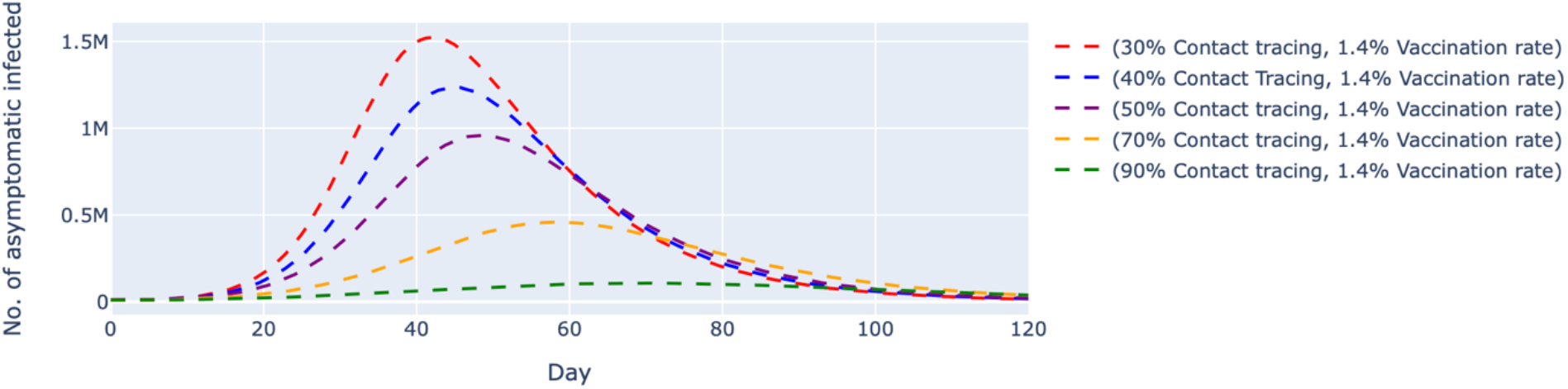
Simulated number of untraced, COVID-19 asymptomatic cases with fixed vaccination rate and varied contact tracing effectiveness. The vaccination rate per day was fixed at 1.4%, which translates to approximately 450,000 doses of vaccine, and the contact tracing effectiveness varied between 30% and 90%. The dashed lines represent the simulated number of untraced COVID-19 asymptomatic cases for five different contexts over 120 days.

**Figure 2b:**
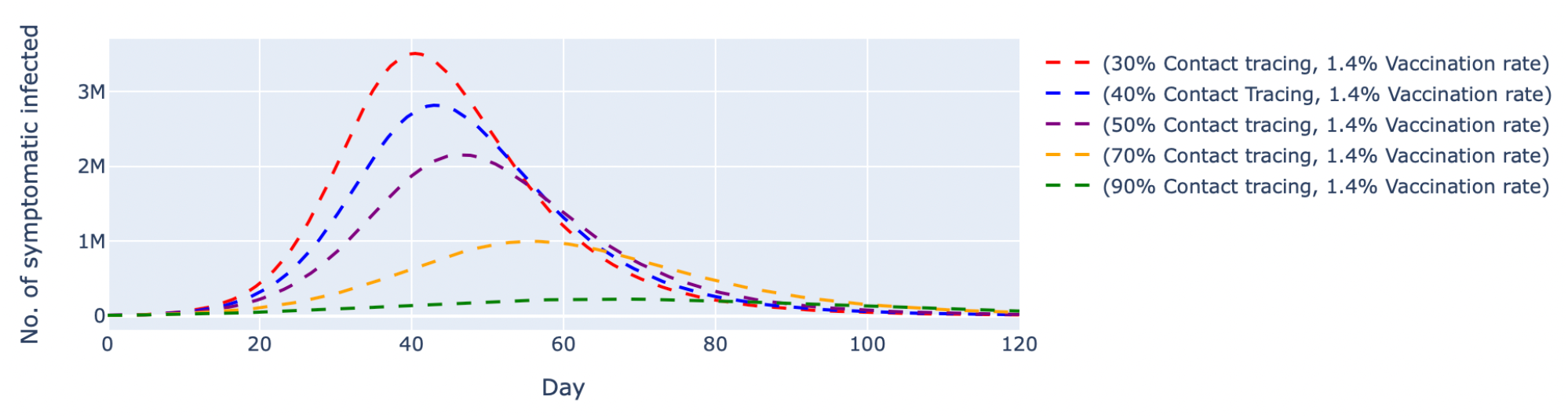
Simulated number of untraced, COVID-19 symptomatic cases with fixed vaccination rate and varied contact tracing effectiveness. The vaccination rate per day was fixed at 1.4%, which translates to approximately 450,000 doses of vaccine, and the contact tracing effectiveness varied between 30% and 90%. The dashed lines represent the simulated number of untraced COVID-19 symptomatic cases for five different contexts over 120 days.

In scenario 2, we conducted simulations by pairing higher contact tracing effectiveness with a lower daily vaccination rate and vice versa. Despite a higher vaccination rate, the simulated trend of new daily untraced, asymptomatic cases contingent on 30% contact tracing effectiveness was estimated to peak at day-42 with 1.46 million cases. The variant with 90% contact tracing effectiveness but a lower vaccination rate delayed peaking at day-74 with 182,000 cases (**Figure 3a**). The same trends were observed among symptomatic cases (**Figure 3b**). A low contact tracing effectiveness of 30% with a high daily vaccinate rate of 1.5% would lead to a peak at 3.35 million untraced, symptomatic cases on day-41. The scenario with the combination of 90% contact tracing effectiveness and only 1% vaccination rate managed to delay the peak to day-72 with 381,000 cases.

**Figure 3a:**
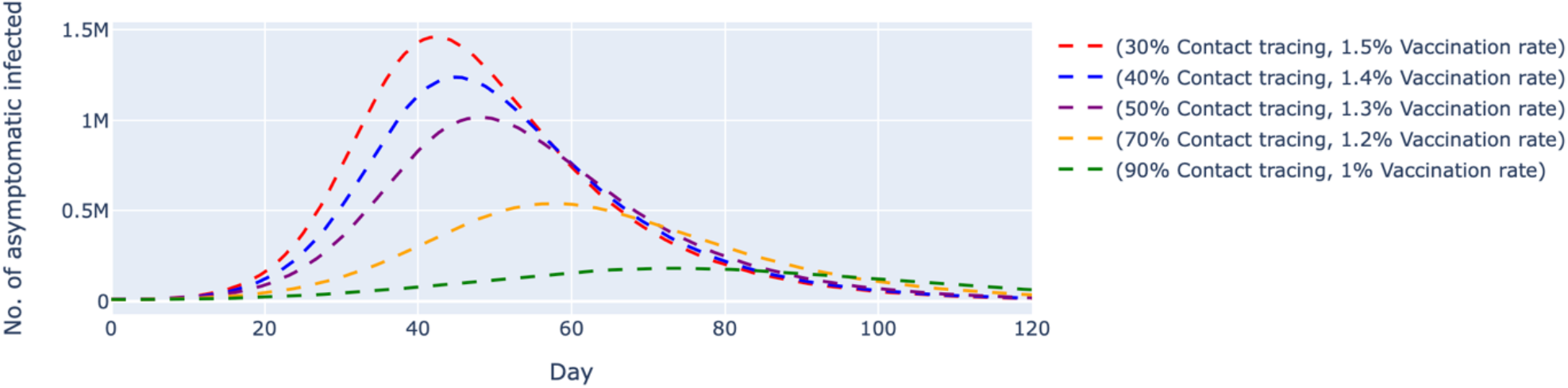
Simulated number of untraced, COVID-19 asymptomatic cases with varied vaccination rates and contact tracing effectiveness. The vaccination rates per day were varied between 1% and 1.5%, which translates to approximately 320,000 and 480,000 doses of vaccine, respectively. The contact tracing effectiveness varied between 30% and 90%. The dashed lines represent the simulated number of untraced COVID-19 asymptomatic cases for five different contexts over 120 days.

**Figure 3b:**
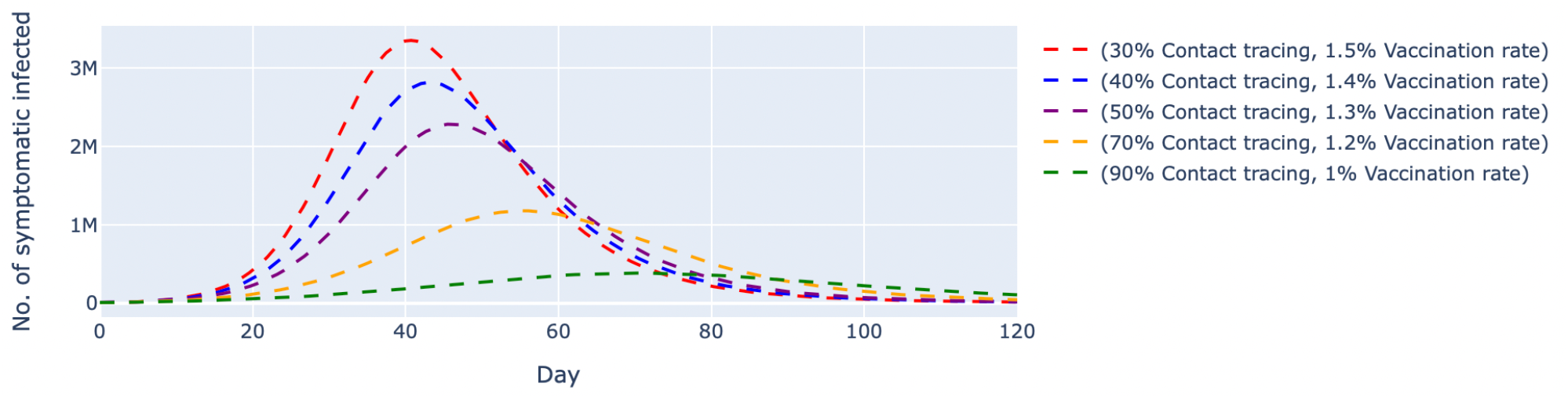
Simulated number of untraced, COVID-19 symptomatic cases with varied vaccination rates and contact tracing effectiveness. The vaccination rates per day were varied between 1% and 1.5%, which translates to approximately 320,000 and 480,000 doses of vaccine, respectively. The contact tracing effectiveness varied between 30% and 90%. The dashed lines represent the simulated number of untraced COVID-19 symptomatic cases for five different contexts over 120 days.

We also simulated two other scenarios where the contact tracing effectiveness was fixed at 90% (**Figure 4a-4b**) and 35% (**Figure 5a-5b**), respectively. In both scenarios, the vaccination rates varied between 0.8% and 1.4% per day. At 90% contact tracing effectiveness (**Figure 4a**), the peaks for all permutations occurred at around the same time (day-68 to day-73) and ranged between 107,000 to 207,000 cases. The estimated highest number of daily untraced, asymptomatic cases differed by approximately 100,000 between high and low vaccination rates when contact tracing effectiveness was fixed. The increase in vaccination rate could only delay the peak but failed to lower the peak significantly. For the untraced, symptomatic cases in **Figure 4b**, a combination of low vaccination rate (1%) and high contact tracing effectiveness (90%) could successfully reduce the highest number of daily cases to about 381,000 at day-70. Under the same circumstance except for a lower contact tracing effectiveness (**Figure 5a**), the estimated number of daily untraced, asymptomatic cases was at 540,000 at day-57 when the vaccination rate was 1.4%. A similar trend could be seen for the untraced, symptomatic cases in **Figure 5b**, whereby a high vaccination rate (1.4%) but low contact tracing effectiveness (35%) could still cause the peak of cases to be 1.2 million at day-54.

**Figure 4a:**
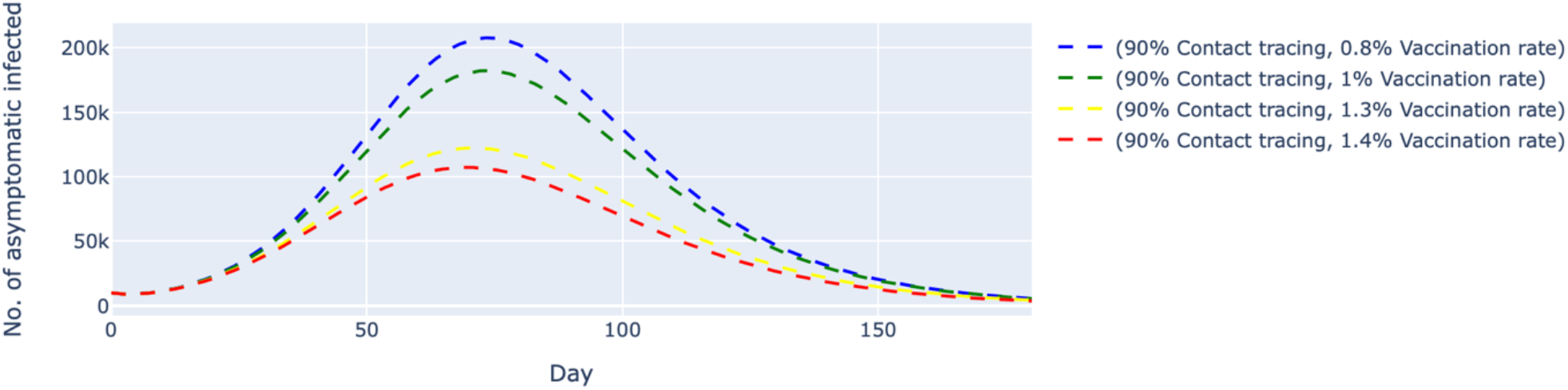
Simulated number of untraced, COVID-19 asymptomatic cases with fixed contact tracing effectiveness at 90% and varied vaccination rates. The contact tracing effectiveness was fixed at 90% and the vaccination rates per day were varied between 0.8% and 1.4%, which translates to approximately 260,000 and 450,000 doses of vaccine, respectively. The dashed lines represent the simulated number of untraced COVID-19 asymptomatic cases for five different contexts over 160 days.

**Figure 4b:**
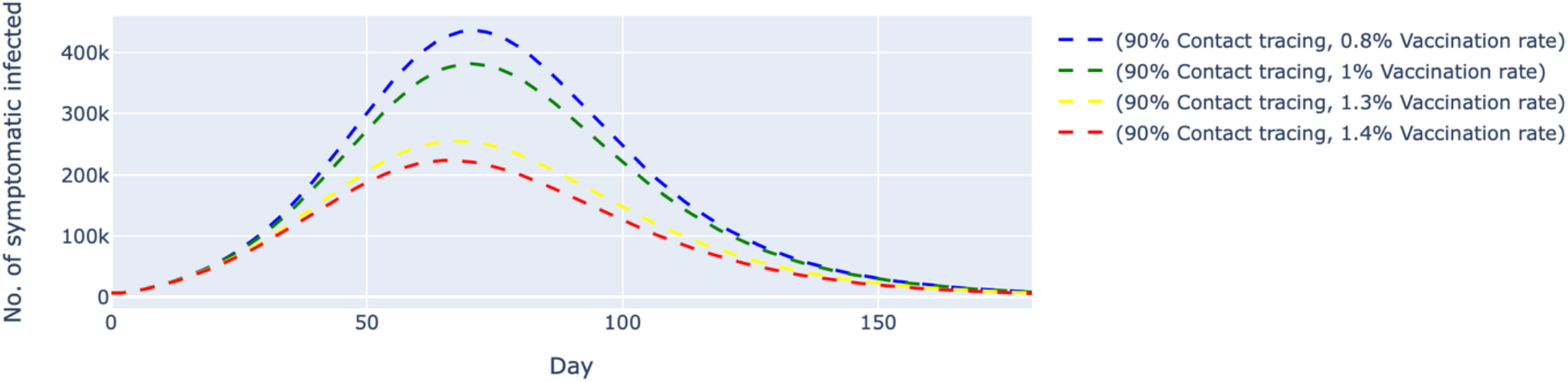
Simulated number of untraced, COVID-19 symptomatic cases with fixed contact tracing effectiveness at 90% and varied vaccination rates. The contact tracing effectiveness was fixed at 90% and the vaccination rates per day were varied between 0.8% and 1.4%, which translates to approximately 260,000 and 450,000 doses of vaccine, respectively. The dashed lines represent the simulated number of untraced COVID-19 symptomatic cases for five different contexts over 160 days.

**Figure 5a:**
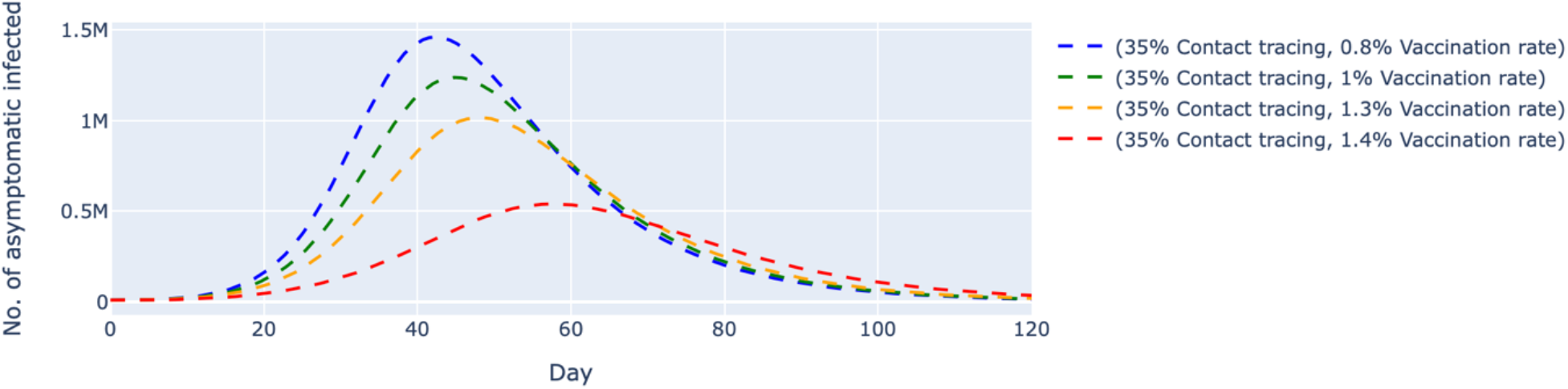
Simulated number of untraced, COVID-19 asymptomatic cases with fixed contact tracing effectiveness at 35% and varied vaccination rates. The contact tracing effectiveness was fixed at 35%, and the vaccination rates per day were varied between 0.8% and 1.4%, which translates to approximately 260,000 and 450,000 doses of vaccine, respectively. The dashed lines represent the simulated number of untraced COVID-19 asymptomatic cases for five different contexts over 120 days.

**Figure 5b:**
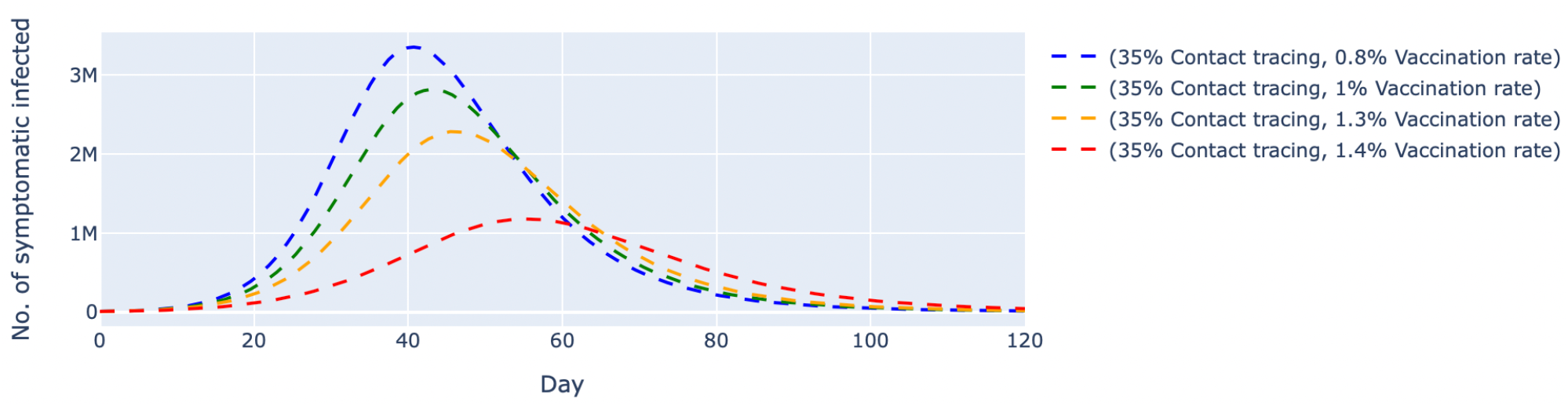
Simulated number of untraced, COVID-19 symptomatic cases with fixed contact tracing effectiveness at 35% and varied vaccination rates. The contact tracing effectiveness was fixed at 35%, and the vaccination rates per day were varied between 0.8% and 1.4%, which translates to approximately 260,000 and 450,000 doses of vaccine, respectively. The dashed lines represent the simulated number of untraced COVID-19 symptomatic cases for five different contexts over 120 days.

## Discussion

The pandemic saw major movement and travel restrictions, lockdown, and personal protective measures implemented globally in various forms and stringency. The Public Health and Social Measures (PHSM) are effective in limiting COVID-19 transmission and death.^24^ However, some of the interventions, particularly lockdowns and cessation of economic activities, have negatively impacted the economy and psychosocial well-being of the affected populations.^25,26^ One major component of PHSM is the enhancement of surveillance and response actions through contact tracing, testing and isolating close contacts, and providing necessary support mechanisms.^24^

In this paper, we simulated the impact of contact tracing on COVID-19 transmission and infection in the context of rising vaccination rates. We observed that a combination strategy of high daily vaccination rate and low contact tracing effectiveness would significantly increase the untraced infectious population. While the vaccine has been poised as one of the most vital tools to take us towards the restoration of post-pandemic normality, we found that contact tracing is key to COVID-19 control. However, the ability of countries to perform contact tracing is challenged by the lack of human resources (contact tracers), compliance to self-isolation orders, and a paucity of timely and accurate contacts data. Successes observed in countries like Singapore and South Korea leveraged technology to aid contact tracing,^6^ and integration of digital technology and the conventional contact tracing approach could result in a swifter response to stem COVID-19 transmission.

In Malaysia, personal details are collected for entry to all premises outside of residence using a mobile application (MySejahtera®) or documented in writing for contact tracing purposes.^28,29^ However, contact tracing of private social gathering at homes remains an issue and has to be carried out manually – therefore hampering the effectiveness of contact tracing. With the rapid rise in COVID-19 infections and an overwhelmed public health system, the contact tracing effectiveness in Malaysia has crippled and currently stands at approximately 30% to 40%.^21,22^ Given the significance of contact tracing in pandemic control, it is pivotal to capitalize the contacts data, automate the data processing and application process,^4^ and develop more efficient strategies to improve contact tracing effectiveness.

In this study, we modeled the number of untraced cases, both symptomatic and asymptomatic. Since this population contributes to forward transmission, an uncontrolled number of these untraced and un-isolated individuals could potentially cause a surge in cases COVID-19 cases. In addition to transmission and infection, future work could be considered using an extended model that incorporates disease severities and health system capacity to estimate the effect of contact tracing on COVID-19 mortality. Furthermore, we set the two main intervention parameters—contact tracing and vaccine—to approximate the ground realities in Malaysia. Notwithstanding testing only on these two PHSMs, we observed the scenario with low contact tracing and increasing vaccination rates successfully mimicked the current transmission trend in Malaysia. However, further parameterization using local data is warranted to generate outcome estimates more salient to Malaysia.

Contact tracing has been at the forefront in controlling the spread of infectious diseases, and an effective and efficient system could prevent disease spread, save lives, and allow the economy to resume.^27^ While vaccination rates have progressively increased in Malaysia and some parts of the world, efficient contact tracing must be rapidly implemented to reach, find, test, isolate, and support the affected populations to bring the pandemic under control.

## Data Availability

Malaysian Covid-19 vaccination data can be found in the national JKJAV website.

https://www.vaksincovid.gov.my

## Notes

### Competing Interest Statement

The authors have declared no competing interest.

### Funding Statement

The work was funded by AINQA Health, Malaysia.

